# The effects of economic difficulties on social and health care costs of children –target trial emulation using complete birth cohort data in Finland

**DOI:** 10.1101/2023.08.11.23293974

**Authors:** Aapo Hiilamo, Markus Keski-Säntti, Aapo Juutinen, Lauri Mäkinen, Tiina Ristikari, Tea Lallukka

**Affiliations:** Itla Children’s Foundation, Helsinki, Finland; Finnish Institute for Health and Welfare, Helsinki, Finland; Department of Public Health, Faculty of Medicine, University of Helsinki, Helsinki, Finland

## Abstract

It is unclear how much costs economic difficulties incur to the health and social care sector, which is a critical research gap to support the economic case for preventing child poverty. We examine the health and social service costs due to families entering into, and transitioning out of, social assistance used as a proxy measure for economic difficulties. We analyzed register data on all Finnish children born in 1997 in the framework of a non-randomized target trial. Inverse probability treatment weighting techniques were used to make the comparison group similar to the treatment group in terms of 29 pretreatment variables. Entry to social assistance was associated with some 1398-2591€ (50%) higher cumulative health and social care costs of the children three years after their families transitioned to social assistance, compared to the group that did not enter to social assistance system. This difference was primarily attributed to higher social care costs. Continued social assistance use was associated with some 1018-2775€ (31%) higher costs compared to the comparison group that exited social assistance. These findings support an economic argument to prevent families from entering economic difficulties and to help those in such situations to transition out.

## Background

In 2021, one-fourth of all children in the EU were at risk of poverty or social exclusion, while the rates varied significantly among the member countries (1). The cost-of-living crises that emerged in 2022 have likely exacerbated the economic difficulties faced by families with children (2). Child poverty is associated with poorer health outcomes and an increased need for social services (3), making the reduction of economic difficulties in families with children a public health priority.

A systematic review of 54 quasi-experimental studies concluded that child poverty has a robust association with a range of negative outcomes (3). Economic difficulties are linked to an increased risk of child mental health problems (4–6), poorer physical health (7), maltreatment (8,9), chronic stress (10) and behavioural problems (11,12). Exogenously introduced changes in economic circumstances, such as social policy programs (13,14) or economic recessions (15), link to changes in these outcomes, supporting the claim that a causal link exists between the two. The key mechanisms between economic difficulties and health problems include lack of access to health care, psychological stress, shame and the social exclusion arising therein, health behaviour, living conditions, and health literacy (5).

While the cost of the human suffering attributed to child poverty cannot fully be quantified, the existing attempts to calculate its monetary impact on the public rely dominantly on expert opinions. One study estimated that child poverty costs approximately 1.03 trillion dollars annually in the U.S., with ill health accounting for 19 per cent and child maltreatment 4 per cent of the total cost (16). Another investigation suggested a smaller annual cost of 500 billion dollars (17). The OECD has posited that childhood socioeconomic disadvantage, a broader manifestation of economic deprivation, has a total monetary impact equivalent to 1.4 per cent of GDP in terms of its later life health effects, and 3.4 per cent of GDP in terms of total effects (18). Although these analyses offer valuable insights into the magnitude of the costs linked to economic difficulties experienced by children, their calculations often lack specificity and unclear counterfactuals to which costs are compared. Some observational studies with individual level data have found, that food insecurity links to higher health expenditures (19) and that low parental income links to higher health care costs (20). But, in our literature search, we did not identify any observational studies that used a target trial framework to assess the costs of childhood economic difficulties. Target trial emulation is a conceptual tool to help researchers to formulate their research questions and their parameters of interest in a straightforward fashion. The target trial design mimics a randomized trial when such trials are not feasible due to practical or ethical concerns. In this framework an ideal trial is first conceptualized by defining the treatment and comparison groups, the start of the idealized trial and its follow-up period and the outcomes of interest (21). Statistical techniques, such as propensity score weighting, are then used to make the distribution of key confounders similar between the treated and comparison groups (22).

There is a need for a robust assessment of the public savings that could be achieved by reducing economic difficulties in families. Such research helps policymakers to allocate resources aimed at mitigating the impacts of economic difficulties in families. In this study, we take advantage of the target trial framework and calculate the health and social care costs attributed to economic difficulties in families with children. The target trial framework is particularly useful in assessing the economic burden of child poverty because it provides a clear counterfactual and avoids common pitfalls with unclear parameters of interest (21), which is often a problem in studies on the association between economic difficulties and negative outcomes. We use social assistance recipiency as a proxy measure for economic difficulties. We focus on the cost of children in the families transitioning to social assistance.

## Methods

We follow the STrengthening the Reporting of OBservational studies in Epidemiology guidelines for observational cohort studies and use the Finnish birth cohort data of 1997 (FB1997). FB1997 is a cohort study consisting of all children born in Finland in 1997. The cohort is constructed using the unique personal number assigned to all residents in Finland. All individuals born in Finland in 1997 were identified from the national birth register and linked to their parents. Both parents and their children were linked to a range of administrative register data using the personal identification numbers. The dataset and its key findings are elaborated in previous studies (23). The ethical approval for the use of this dataset has been obtained from the Ethics Committee of the Finnish Institute for Health and Welfare, THL (§572/2013). For the study at hand, we use register data drawn from the healthcare treatment notification register maintained by the THL, the population register by the Digital and Population Data Services Agency, the Social assistance register, and educational registers managed by Statistics Finland, among others.

The original birth cohort consisted of 58 802 children and their parents (mother n= 57 888 and father n= 57 222), from which we excluded individuals who moved abroad, deceased and children not living with their parents. Furthermore, we dropped randomly all but one of the siblings who had the same parents and thus had identical parental data. Our unit of analysis is children-year while we measure a number of variables at the child and family levels. The two target trials (see details below) had 54 858 and 16 026 cohort members including 697 680 and 71 131 children-years in total.

## Treatment

Our treatment variable is a change in the recipiency of social assistance of the family in which the children lived. Social assistance is granted and measured at the family level. The data is derived from the national social assistance register. Data on social assistance recipiency was originally gathered from local municipalities and is managed by the Finnish Institute for Health and Welfare. We define social assistance recipiency as any situation where children’s parents had received social assistance at least once within the given calendar year. When a parent did not reside with their children (for example, due to a divorce), their social assistance records are not considered in our analysis.

In line with our previous research with the same birth cohort data (24,25), we use social assistance as a proxy measure for economic difficulties in the family. Social assistance is intended for all households in Finland who have no spare income or assets to cover some basic living costs. For instance, a single adult was entitled to approximately 555 euros monthly in 2023. Other costs such as housing, medical care, and certain essentials are covered on a means-tested evaluation. Some tenth of children in Finland lives in a household that has received social assistance at least once in 2021 (26).

## Outcome

Our outcome variable of interest is the costs of specialized health care, social care and prescribed medication reimbursements in 2018 prices. In calculating the costs of specialized medical care, we used the Nordic Diagnosis Related Groups classification (NordDRG). NordDRG is a patient classification system used in Nordic countries that can be used to estimate the costs of specialized medical care (27). However, the NordDRG classification is not well suited for estimating the costs of psychiatric specialities. Thus, separate cost estimates have been made for them using appropriate daily prices extrapolated from the THL’s reports on the Unit Costs of Health and Social Care in Finland in 2001 (28) and Unit Costs of Health and Social Care in Finland in 2011 (29). Costs in social care were calculated based on the daily rates of out-of-home care extrapolated from the report on the Comparison of Child Protection Services and Costs in the Six Largest Cities in Finland in 2015 (30).

Data on social care costs are based on the Register of Child Welfare (31). Register of Child Welfare includes data from the grounds and duration of placement outside the home and the social care costs can be estimated by multiplying the duration by the appropriate daily rate. However, the Register of Child Welfare does not include information about open care and therefore these costs are not included in the analysis. Medicine reimbursement costs are based on the defined daily dose (DDD), which is a statistical measure of drug consumption. Data on medication reimbursements are from the registers of the Social Insurance Institution of Finland. Our data includes only psycholeptic and psychoanaleptic drugs. Costs of medicine reimbursement are estimated by multiplying the DDD included in the register by an average cost of one DDD extrapolated from the Finnish Statistics on Medicines (32).

## Control variables

To control for confounding, we derived 29 variables from administrative registers that were measured before the treatment. In our probability of treatment model, we included parental health status, measured using records of specialized health care records (four variables), unemployment status (two variables), highest educational attainment (two variables), and income, transformed logarithmically for both the mother and father. Dichotomous variables accounting for zero income were included to measure extreme financial difficulties. We included variables measuring previous cumulative time spent in social assistance. We included cohort members’ sex, birth weight, imputed birth weight, and current values of social, psychiatric, and home care costs. We also calculated cumulative sums of these costs before the treatment and transformed them logarithmically to account for potential non-linear relationships.

## The definitions of the target trials

We use the target trial framework (21). We defined two types of target trials. The first is labelled as the “transition to social assistance use” trial. The target population in this trial is all children in families not receiving social assistance in a given calendar year (t). The intervention group includes children in families that transition to receiving social assistance in the subsequent year (t+1). This group is compared with the comparison group, which consists of children in families that continue without social assistance in the following year (t+1). The outcome measure of the trial is the health and social care costs incurred for these children two to four years later. By examining how this transition is associated with subsequent health and social care costs, we aim to assess the potential long-term effects of preventing economic difficulties in families with children on children’s health and welfare outcomes.

In the second “continued social assistance use” trial, the population of interest consists of children in families receiving social assistance in a given calendar year (t). The intervention is defined as families receiving social assistance also in the subsequent year (t+1). This group is compared with families that did not receive social assistance in the following year, i.e. transitioned out from social assistance use (t+1). The outcomes are identical to the first trial. This second trial seeks to quantify the potential impacts of interventions helping families with children exit from economic difficulties on the health and welfare costs of children.

We conducted these trials for three populations; first, all children (all years pooled), to test moderation by age, children aged 0-7 before the change in social assistance status (years 1998–2004 pooled) and children aged 8-14 (2004–2011).

## Statistical analysis

### Parameters of interest

Our parameter of interest is the Average Treatment Effect on the Treated (ATT). We selected ATT as our primary estimand due to its policy relevance. ATT provides information about the potential savings to be achieved if the transition into social assistance usage could be completely prevented. For the first trial - the transition to social assistance use - we are contrasting a counterfactual scenario in which families did not transition into social assistance, with the real-world scenario in which these same families did. This is to say that ATT quantifies the mean difference between the counterfactual and actual scenario. Thus, the population who transitioned in real life to social assistance is the reference population to which the comparison group is made similar to. In the second trial – the “continued social assistance use” trial - we are comparing a hypothetical scenario where families did transition out from social assistance, with the actual scenario, where these families continued to receive social assistance. ATT in this trial quantifies the potential savings in costs that could result if the families that were unsuccessful in transitioning out from social assistance would have been successful.

### Estimation

To estimate these parameters, we used inverse probability treatment weighting. We start by estimating the probability of treatment models in which we predicted the probability of social assistance status at t+1 separately for each trial. We fit logistic regression models regressing the social assistance status at time t+1 on the 29 covariates listed above. We fit these models by year to allow all interactions with a year of measurement. From the model, we then predict propensity scores (PS) and set the inverse probability of treatment weights to 1 if the observation was in the treatment group and as 1/(1-ps) if the observation was in the comparison group. We then calculate the weighted averages of the costs in the social and health care registers in the following calendar years from the social assistance transition. We present the balance statistics as recommended (33). To assess the uncertainty of our estimates, we use the bootstrapping method and calculate 95 percentile intervals from 500 replications. Our estimation allows us to infer causality under assumptions of no misspecifications in the treatment model and no unmeasured confounding over and above the dimensions captured by the measured control variables (34). To assess the potential of model misspecifications, we ran regression model estimates as a supplementary analysis. R software version 4.1.1. was used in all phases of the analysis.

## Results

The trial investigating the transition to social assistance use consisted of 680,314 observations in the comparison group and 17,387 in the treatment groups; that is, they transitioned to social assistance use. The children in the treated group had a lower parental education level, more parental health records, a higher share of parental unemployment, and lower parental income compared to the comparison group who did not transition to social assistance (Table 1). Previous social and health care costs of children in the treatment group were also higher. However, IPTW balanced these differences, and few variables had standardized differences higher than 0.1 (Figure 1).

**Figure 1.**
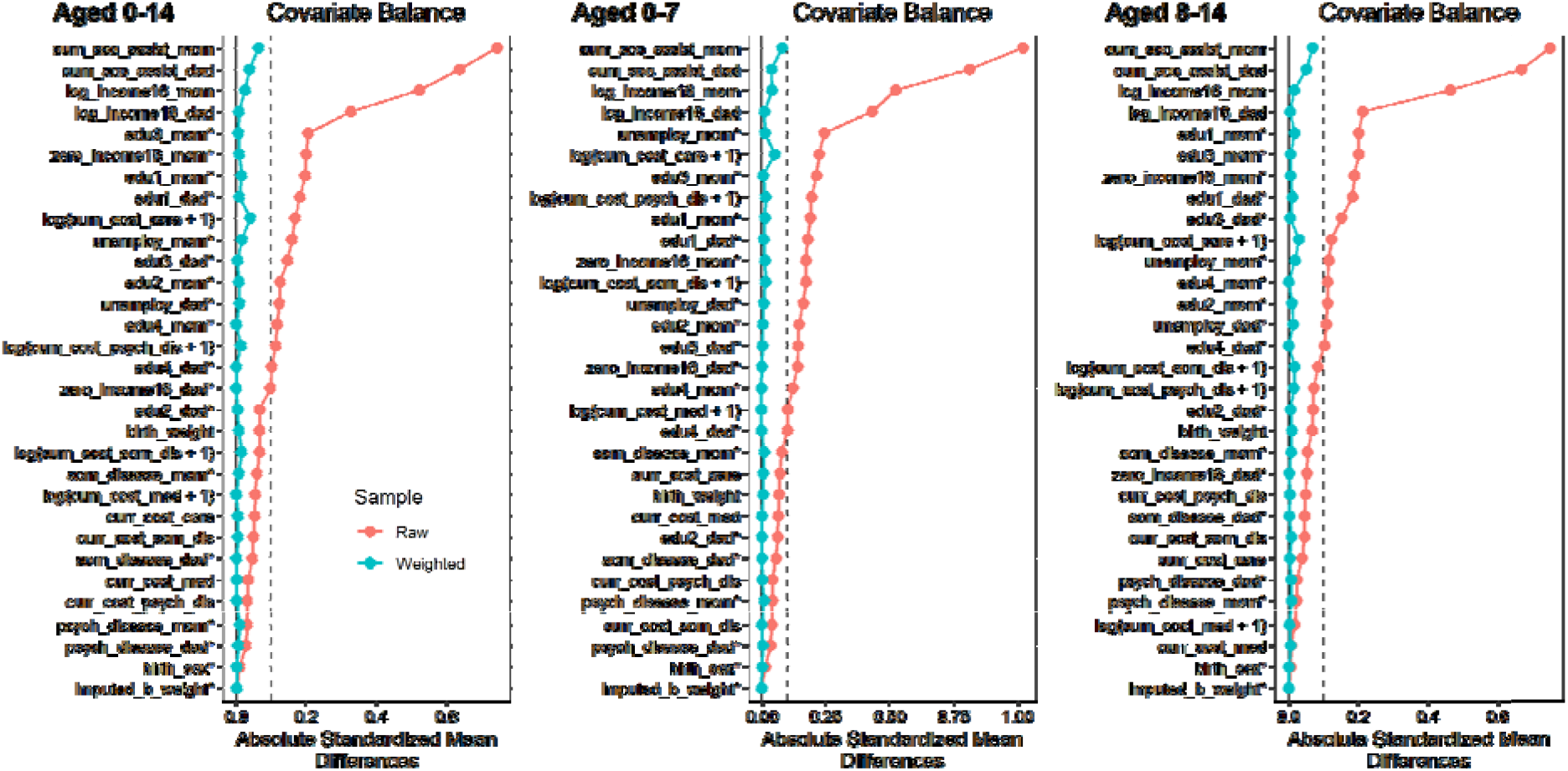
Balance characteristics of the study population in the social assistance entry trial. Absolute mean differences before and after weighting. observations = 697,680 The Finnish Birth Cohort 1997.

**Table 1.** Characteristics of the study population in the social assistance entry trial. Treated group are children in families entering to social assistance use. The comparison group are children in families not entering to social assistance use. The Finnish Birth Cohort 1997.

|  | All children |  | Children aged 0-7 |  | Children aged 8-14 |  |
| --- | --- | --- | --- | --- | --- | --- |
|  | Comparison | Treated | Comparison | Treated | Comparison | Treated |
| *Mother's education* |  |  |  |  |  |  |
| Lower university level | 36.85 % | 16.28 % | 37.01 % | 17.04 % | 36.71 % | 15.22 % |
| Middle level | 40.42 % | 52.95 % | 40.15 % | 51.23 % | 40.68 % | 55.37 % |
| Basic level | 9.27 % | 29.03 % | 10.04 % | 30.03 % | 8.54 % | 27.62 % |
| At least upper university level | 13.46 % | 1.74 % | 12.8 % | 1.7 % | 14.07 % | 1.79 % |
| Father's education level |  |  |  |  |  |  |
| Lower university level | 25.48 % | 10.8 % | 25.72 % | 10.66 % | 25.26 % | 10.99 % |
| Middle level | 46.55 % | 53.28 % | 46.47 % | 53.46 % | 46.62 % | 53.03 % |
| Basic level | 15.65 % | 33.84 % | 15.65 % | 33.95 % | 15.66 % | 33.69 % |
| At least upper university level | 12.31 % | 2.08 % | 12.16 % | 1.93 % | 12.46 % | 2.29 % |
| Mother's somatic illness | 21.51 % | 27.48 % | 19.23 % | 24.52 % | 23.65 % | 31.65 % |
| Father's somatic illness | 18.87 % | 23.56 % | 16.86 % | 21.45 % | 20.75 % | 26.52 % |
| Father's unemployment | 6.87 % | 19.16 % | 2.78 % | 13.56 % | 10.7 % | 27.03 % |
| Mother's unemployment | 9.82 % | 25.75 % | 4.94 % | 16.41 % | 14.4 % | 38.9 % |
| Father's psychiatric illness | 1.19 % | 3.9 % | 0.83 % | 3.15 % | 1.53 % | 4.95 % |
| Mother's psychiatric illness | 1.43 % | 4.49 % | 0.99 % | 3.31 % | 1.85 % | 6.16 % |
| Birth weight imputed* | 0.22 % | 0.12 % | 0.22 % | 0.13 % | 0.22 % | 0.1 % |
| Cumulative somatic health care costs | 3960.62 | 4554.73 | 2918.99 | 3390.64 | 4936.05 | 6193.32 |
| Cumulative psychiatric health care costs | 316.14 | 628.66 | 109.15 | 173.08 | 509.97 | 1269.93 |
| Cumulative social care costs | 210.16 | 1448.88 | 58.53 | 467 | 352.14 | 2830.97 |
| Father's income (log) | 9.01 | 7.61 | 8.47 | 7.58 | 9.51 | 7.66 |
| Mother's income (log) | 8.09 | 5.72 | 6.97 | 4.86 | 9.15 | 6.91 |
| No income father | 0.13 | 0.23 | 0.18 | 0.23 | 0.09 | 0.23 |
| No income mother | 0.18 | 0.38 | 0.27 | 0.46 | 0.09 | 0.27 |
|  | Comparison | Treated | Comparison | Treated | Comparison | Treated |
| Previous time in social assistance mother | 0.52 | 2.73 | 0.28 | 1.59 | 0.75 | 4.35 |
| Previous time in social assistance father | 0.5 | 2.37 | 0.26 | 1.43 | 0.72 | 3.68 |
| Birth weight | 3558.92 | 3521.38 | 3560.57 | 3523.04 | 3557.39 | 3519.05 |
| Social care costs before treatment | 36.9 | 215.22 | 9.75 | 73.81 | 62.33 | 414.26 |
| Psychiatric health care costs before treatment | 61.52 | 159.79 | 33.77 | 65.46 | 87.5 | 292.57 |
| Somatic health care costs before treatment | 346.22 | 472.27 | 429.58 | 562.67 | 268.15 | 345.03 |
| Medicine costs before treatment | 1.67 | 3.06 | 0.05 | 0.06 | 3.19 | 7.28 |
| n | 680314 | 17366 | 328992 | 10153 | 351322 | 7213 |

\* This is binary variable that birth weight data was not available and imputed for the mean of the sample.

After balancing, the two groups had little difference in the costs in the year before the transition (a t-1 difference of -23€ [bootstrap 95% percentile interval of -431–32]) but had significant differences a year (608€ [406–762]), two years (742€ [510–943]), and three years after the transition (742€ [507–997], Table 2). The cumulative three-year difference was 2092€ per child (1511–2619€). This is to say that the treatment group had some 50 per cent higher costs than the comparison group. This difference was due to higher social care costs (1794€ [1276–2260] difference, 72% higher than the comparison group). Smaller differences were observed in somatic health care costs (151€ [36–263] 14% difference) in psychiatric health care costs in the treatment group (146€ [4–316], 23% difference). Age-stratified analysis suggested that the reduction was stronger in absolute terms in the 8-14 aged population (a 3372€ [2108–4405] difference) than in the 0-7 aged population (a 1185€ [774–1599] difference).

**Table 2.** Costs estimates in Euros for the social assistance entry trial, estimated via inverse probability treatment weighting. ATT is the average treatment effect on the treated. Treated group are children in families entering to social assistance use. The comparison group are children in families not entering to social assistance use. Observations = 697680. The Finnish Birth Cohort of 1997.

|  | **ALL children** |  | **Children aged 0-7 (aged 1-7)** |  | **Children aged 8-14 (8-14)** |  |
| --- | --- | --- | --- | --- | --- | --- |
|  | ATT (95% bootstrap percentile) | ATT/comparison mean | ATT (95% bootstrap percentile) | ATT/comparison mean | ATT (95% bootstrap percentile) | ATT/comparison mean |
| All costs |  |  |  |  |  |  |
| T-1 | -23 [-431-32] | -3% | -29 [-532-20] | -4% | -15 [-379-97] | -1% |
| T+1 | 608 [406-762] | 56% | 421 [244-570] | 70% | 873 [499-1196] | 49% |
| T+2 | 742 [510-943] | 52% | 403 [243-564] | 60% | 1221 [763-1648] | 49% |
| T+3 | 742 [507-997] | 44% | 361 [184-540] | 46% | 1279 [733-1749] | 43% |
| Sum t1-t3 | 2092 [1511-2619] | 50% | 1185 [774-1599] | 58% | 3372 [2108-4405] | 46% |
| Health care costs - somatic |  |  |  |  |  |  |
| T-1 | -13 [-395-9] | -3% | -22 [-487-11] | -4% | 0 [-84-22] | 0% |
| T+1 | 66 [4-114] | 18% | 112 [21-190] | 30% | 1 [-43-33] | 0% |
| T+2 | 56 [8-107] | 16% | 76 [12-148] | 22% | 28 [-18-79] | 7% |
| T+3 | 29 [-6-64] | 8% | 56 [8-106] | 17% | -10 [-48-31] | -2% |
| Sum t1-t3 | 151 [36-263] | 14% | 244 [54-405] | 23% | 19 [-75-110] | 2% |
| Health care costs - psychiatric |  |  |  |  |  |  |
| T-1 | 6 [-47-43] | 4% | -2 [-13-5] | -3% | 19 [-249-101] | 7% |
| T+1 | 47 [-26-113] | 24% | -12 [-91-42] | -10% | 130 [10-289] | 42% |
| T+2 | 62 [-1-134] | 30% | 5 [-46-47] | 5% | 143 [-3-309] | 41% |
| T+3 | 38 [-44-157] | 17% | 0 [-37-28] | 0% | 91 [-112-323] | 24% |
| Sum t1-t3 | 146 [4-316] | 23% | -8 [-110-82] | -2% | 364 [16-702] | 35% |
| Social care costs |  |  |  |  |  |  |
| T-1 | -17 [-85-16] | -7% | -5 [-74-25] | -6% | -34 [-114-28] | -8% |
| T+1 | 496 [335-621] | 96% | 321 [205-418] | 300% | 742 [420-1016] | 68% |
| T+2 | 624 [430-794] | 73% | 322 [196-439] | 147% | 1049 [630-1423] | 60% |
| T+3 | 675 [456-878] | 61% | 305 [159-463] | 91% | 1196 [756-1641] | 54% |
| Sum t1-t3 | 1794 [1276-2260] | 72% | 948 [610-1266] | 143% | 2987 [1866-3970] | 59% |
| Medical reimbursement costs |  |  |  |  |  |  |
| T-1 | 0 [0-0] | 0% | 0 [0-0] | NaN% | 0 [0-1] | 0% |
| T+1 | 0 [-1-1] | 0% | 0 [0-0] | 0% | 1 [-1-3] | 8% |
| T+2 | 0 [-1-1] | 0% | 0 [-1-0] | 0% | 1 [-2-3] | 7% |
| T+3 | 1 [-1-2] | 12% | 0 [-1-0] | 0% | 2 [-2-4] | 12% |
| Sum t1-t3 | 1 [-2-4] | 5% | 0 [-2-1] | 0% | 3 [-4-9] | 7% |

The second trial, concerning continued social assistance use, consisted of 22,971 observations in the comparison group and 48,244 in the treatment group, that is, their families continued to use social assistance. The treated group had lower income and longer periods of previous social assistance use than the comparison group that exited social assistance (Table 3). After balancing the comparison group to be similar to the treatment group, there were no significant differences in the costs before the treatment (37 [-176–194] difference). The continued social assistance recipiency group had 518 [130–774] higher costs a year, 636 [302–913] two years, and 820 [475–1136] three years after the treatment, which resulted in a cumulative difference of 1974 [1007–2709], compared to the group transitioning out from social assistance. In relative terms, the treatment group had 31 per cent higher costs. This difference was driven by differences in social care costs (1909 [977–2608], 43%) while small differences were also observed in somatic health care costs (146 [74–216]€, 13%). No significant differences were observed in psychiatric healthcare costs. No clear age group differences were observed due to the uncertainty of the estimates. Supplementary analyses using regression analysis produced similar results.

**Table 3.** Characteristics of the study population in the continued social assistance trial. Treated group are children in families with continued social assistance use. The comparison group are children in families exiting social assistance use. The Finnish Birth Cohort 1997.

|  | **ALL children** |  | **Children aged 0-7** |  | **Children aged 8-14** |  |
| --- | --- | --- | --- | --- | --- | --- |
|  | **Comparison** | **Treated** | **Comparison** | **Treated** | **Comparison** | **Treated** |
| *Mother's education* |  |  |  |  |  |  |
| Lower university level | 17.07 % | 9.08 % | 17.96 % | 9.17 % | 15.37 % | 8.91 % |
| Middle level | 52.96 % | 45.01 % | 51.31 % | 43.42 % | 56.11 % | 48.04 % |
| Basic level | 28.01 % | 44.92 % | 28.72 % | 46.56 % | 26.66 % | 41.81 % |
| At least upper university level | 1.96 % | 0.98 % | 2.01 % | 0.85 % | 1.86 % | 1.24 % |
| Father's education level |  |  |  |  |  |  |
| Lower university level | 10.98 % | 7.6 % | 11.18 % | 7.19 % | 10.6 % | 8.39 % |
| Middle level | 52.67 % | 46.2 % | 52.45 % | 45.98 % | 53.09 % | 46.62 % |
| Basic level | 34.21 % | 45 % | 34.23 % | 45.82 % | 34.17 % | 43.46 % |
| At least upper university level | 2.14 % | 1.19 % | 2.14 % | 1.01 % | 2.15 % | 1.53 % |
| Mother's somatic illness | 26.37 % | 28.41 % | 23.36 % | 25.45 % | 32.1 % | 34.01 % |
| Father's somatic illness | 23.35 % | 25.2 % | 21.39 % | 23.71 % | 27.08 % | 28.01 % |
| Father's unemployment | 26.72 % | 42.95 % | 22.96 % | 42.4 % | 33.85 % | 43.98 % |
| Mother's unemployment | 32.3 % | 39.17 % | 22.96 % | 29.46 % | 50.06 % | 57.53 % |
| Father's psychiatric illness | 4.62 % | 5.77 % | 3.71 % | 5.16 % | 6.34 % | 6.92 % |
| Mother's psychiatric illness | 4.59 % | 6.1 % | 3.29 % | 4.44 % | 7.05 % | 9.25 % |
| Birth weight imputed* | 0.11 % | 0.23 % | 0.13 % | 0.2 % | 0.09 % | 0.28 % |
| Cumulative somatic health care costs | 4349.74 | 4858.36 | 3377.96 | 3739.01 | 6197.55 | 6973.05 |
| Cumulative psychiatric health care costs | 519.91 | 647.44 | 161.68 | 188.12 | 1201.09 | 1515.19 |
| Cumulative social care costs | 1586.3 | 3245.35 | 638.53 | 1522.96 | 3388.46 | 6499.29 |

|  | **ALL**<br>**children** |  | **Children aged**<br>**0-7** |  | **Children aged**<br>**8-14** |  |
| --- | --- | --- | --- | --- | --- | --- |
|  | **Comparison** | **Treated** | **Comparison** | **Treated** | **Comparison** | **Treated** |
| Father's income (log) | 7.45 | 5.73 | 7.45 | 5.72 | 7.46 | 5.75 |
| Mother's income (log) | 5.57 | 3.45 | 4.97 | 2.9 | 6.72 | 4.5 |
| No income father | 0.23 | 0.39 | 0.23 | 0.38 | 0.24 | 0.4 |
| No income mother | 0.38 | 0.6 | 0.43 | 0.65 | 0.27 | 0.49 |
| Previous time in social<br>assistance mother | 4.09 | 5.37 | 3.01 | 3.74 | 6.12 | 8.46 |
| Previous time in social<br>assistance father | 3.22 | 4.45 | 2.45 | 3.21 | 4.67 | 6.79 |
| Birth weight | 3515.53 | 3473.79 | 3516.48 | 3473.53 | 3513.73 | 3474.27 |
| Social care costs before<br>treatment | 318.11 | 706.33 | 175.37 | 428.58 | 589.55 | 1231.06 |
| Psychiatric health care<br>costs before treatment | 123.88 | 151.86 | 55.98 | 65.63 | 252.98 | 314.78 |
| Somatic health care costs<br>before treatment | 529.59 | 575.18 | 627.25 | 674.76 | 343.9 | 387.04 |
| Medicine costs before<br>treatment | 2.39 | 3.34 | 0.08 | 0.09 | 6.78 | 9.5 |
| n | 22971 | 48160 | 15054 | 31491 | 7917 | 16669 |

**Table 4.** Costs estimates in Euros for the continued social assistance use trial, estimated via inverse probability treatment weighting. ATT is the average treatment effect on the treated. Treated group are children in families with continued social assistance use. The comparison group are children in families exiting social assistance use. Observations = 71 131. The Finnish Birth Cohort of 1997.

|  | **ALL children** |  | **Children aged 0-7** |  | **Children aged 8-14** |  |
| --- | --- | --- | --- | --- | --- | --- |
|  | ATT (95% bootstrap percentile to be added) | Treatment/comparison mean | ATT (95% bootstrap percentile to be added) | Treatment/comparison mean | ATT (95% bootstrap percentile to be added) | Treatment/comparison mean |
| All costs |  |  |  |  |  |  |
| T-1 | 37 [-176–194] | 3% | 19 [-151–160] | 2% | 67 [-703–374] | 4% |
| T+1 | 518 [130–774] | 31% | 475 [270–689] | 49% | 589 [-841–1211] | 19% |
| T+2 | 636 [302–913] | 29% | 579 [297–833] | 47% | 728 [-419–1391] | 18% |
| T+3 | 820 [475–1136] | 33% | 723 [413–1000] | 50% | 985 [-140–1691] | 22% |
| Sum t1-t3 | 1974 [1007–2709] | 31% | 1778 [1043–2441] | 49% | 2302 [-1058–4098] | 20% |
| Health care costs - somatic |  |  |  |  |  |  |
| T-1 | 25 [-4–61] | 5% | 27 [-32–67] | 4% | 21 [-24–69] | 6% |
| T+1 | 72 [38–108] | 19% | 86 [41–128] | 23% | 46 [-10–96] | 13% |
| T+2 | 54 [24–86] | 14% | 55 [12–97] | 15% | 51 [0–107] | 13% |
| T+3 | 20 [-17–55] | 5% | 38 [-2–71] | 11% | -14 [-104–70] | -3% |
| Sum t1-t3 | 146 [74–216] | 13% | 180 [78–269] | 17% | 83 [-42–214] | 7% |
| Health care costs - psychiatric |  |  |  |  |  |  |
| T-1 | -5 [-43–30] | -3% | -6 [-25–6] | -8% | -4 [-157–88] | -1% |
| T+1 | -49 [-223–42] | -19% | 3 [-23–26] | 3% | -151 [-601–117] | -28% |
| T+2 | -28 [-110–40] | -11% | -2 [-36–32] | -1% | -78 [-306–106] | -17% |
| T+3 | -3 [-62–48] | -1% | 26 [-6–54] | 19% | -60 [-202–78] | -14% |
| Sum t1-t3 | -80 [-280–82] | -11% | 28 [-42–94] | 7% | -290 [-812–136] | -21% |
| Social care costs |  |  |  |  |  |  |
| T-1 | 17 [-191–151] | 2% | -3 [-145–134] | -1% | 50 [-595–307] | 4% |
| T+1 | 496 [204–716] | 47% | 385 [179–596] | 80% | 695 [-514–1189] | 33% |
| T+2 | 611 [279–868] | 39% | 527 [263–764] | 74% | 757 [-369–1415] | 24% |
| T+3 | 802 [476–1102] | 44% | 659 [368–926] | 68% | 1059 [-39–1687] | 30% |
| Sum t1-t3 | 1909 [977–2608] | 43% | 1571 [863–2254] | 73% | 2511 [-798–4233] | 29% |
| Medical reimbursement costs |  |  |  |  |  |  |
| T-1 | 0 [-2–1] | 0% | 0 [0–0] | NaN% | -1 [-7–2] | -10% |
| T+1 | 0 [-3–1] | 0% | 0 [0–0] | NaN% | -1 [-12–3] | -7% |
| T+2 | -1 [-4-1] | -14% | 0 [-1-0] | 0% | -1 [-14-4] | -6% |
| T+3 | 0 [-3-2] | 0% | 0 [-1-1] | 0% | 0 [-10-5] | 0% |
| Sum<br>t1-t3 | -1 [-10-4] | -5% | 0 [-2-1] | 0% | -2 [-36-11] | -4% |

## Discussion

This study is, to the best of our knowledge, the first population-based investigation to quantify the health and social care costs of children due to economic difficulties in their families using a target trial framework. Our results suggest substantial costs saving potential of preventing economic difficulties in families with children.

In our first mimicked trial, the transition to social assistance, a proxy for economic difficulties in families with children, was associated with an estimated 50 percent increase in health and social care costs per child over a three-year period, compared to the counterfactual of the same group not transitioning to social assistance. A considerable proportion of these costs were due to social care while significant differences, albeit smaller, were observed in health care costs due to other causes. This finding is in line with the long line of research linking entry to economic difficulties with health and development problems of children (3). For example, an UK cohort study showed that transition into income poverty increased risk of child mental health problems (4). Moreover, the cost increase was more substantial in absolute terms among older children than younger children. These findings suggest that the impacts of child poverty may be strong during the later years, but we did not focus on the long-term effects.

Our second target trial focused on the effects of continued social assistance use. We found that children in families that continued in social assistance had 31 percent higher costs compared to the counterfactual of successful transition out social assistance. This result was driven by social care costs. Surprisingly we did not find significant differences in psychiatric health care costs in this trial. This finding that may be related to lasting effects of economic difficulties in families. Nevertheless, the result from this trial highlights the negative effects of persistent economic difficulties on social care costs.

In this study we did not investigate the mechanism through which economic difficulties link to higher costs. Nevertheless, previous studies suggest mechanisms such as increasing mental health problems, family instability and substance abuse, all of which may affect the need for health and social services. To physical health economic difficulties may link through to lack of access to hobbies and healthy everyday resources. Stress may arise from parental economic difficulties which may trigger physical and mental health issues. Economic difficulties may link to stigma, bullying, social exclusion, fewer friends and constrained opportunities for past time activities (5). All of these factors may link to poorer physical health.

While previous estimates suggest that health care is a major economic burden of child poverty (16–18), our results indicate that the main cost concern is social care. This is not surprising considering the accumulated evidence on the link between poverty and child welfare intervention needs. For example, a previous ecological analysis link social assistance use to higher rate of out-of-home care in Finnish municipalities (35).

In interpreting our findings, it should be noted that there are more than one way of transitioning to social assistance or to continue receiving social assistance. Thus, our estimates should be perceived as an unknown weighted average of the various ways in which families transition to social assistance, compared to weighted average of the different ways in which families do not change their social assistance status. Our analyses relied on non-randomized observational data and are thus subject to unmeasured confounding. While we included 29 control variables some residual confounding certainly remains. We did not access to survey data which would have offered more detailed, subjective information useful in controlling for confounding.

Moreover, we used social assistance as a proxy for economic difficulties, which may not always reflect the economic difficulties experienced by families due to stigma of applying for social assistance. Moreover, families not eligible for social assistance may experience financial difficulties, for example, due to significant debt burden.

There are several brackets of costs that were not included in our study. First, we did not have data on primary health care. Second, we lacked information on health care costs of the parents or other family members. Third, we did not make any cost-calculations of specialized services provided in schools. Fourth, we did not take into consideration potential costs in the criminal correction system. Nevertheless, these findings add to increasing body of evidence that support prioritizing efforts to reduce economic difficulties in families as a public health and public saving strategy.

**Figure 1.**
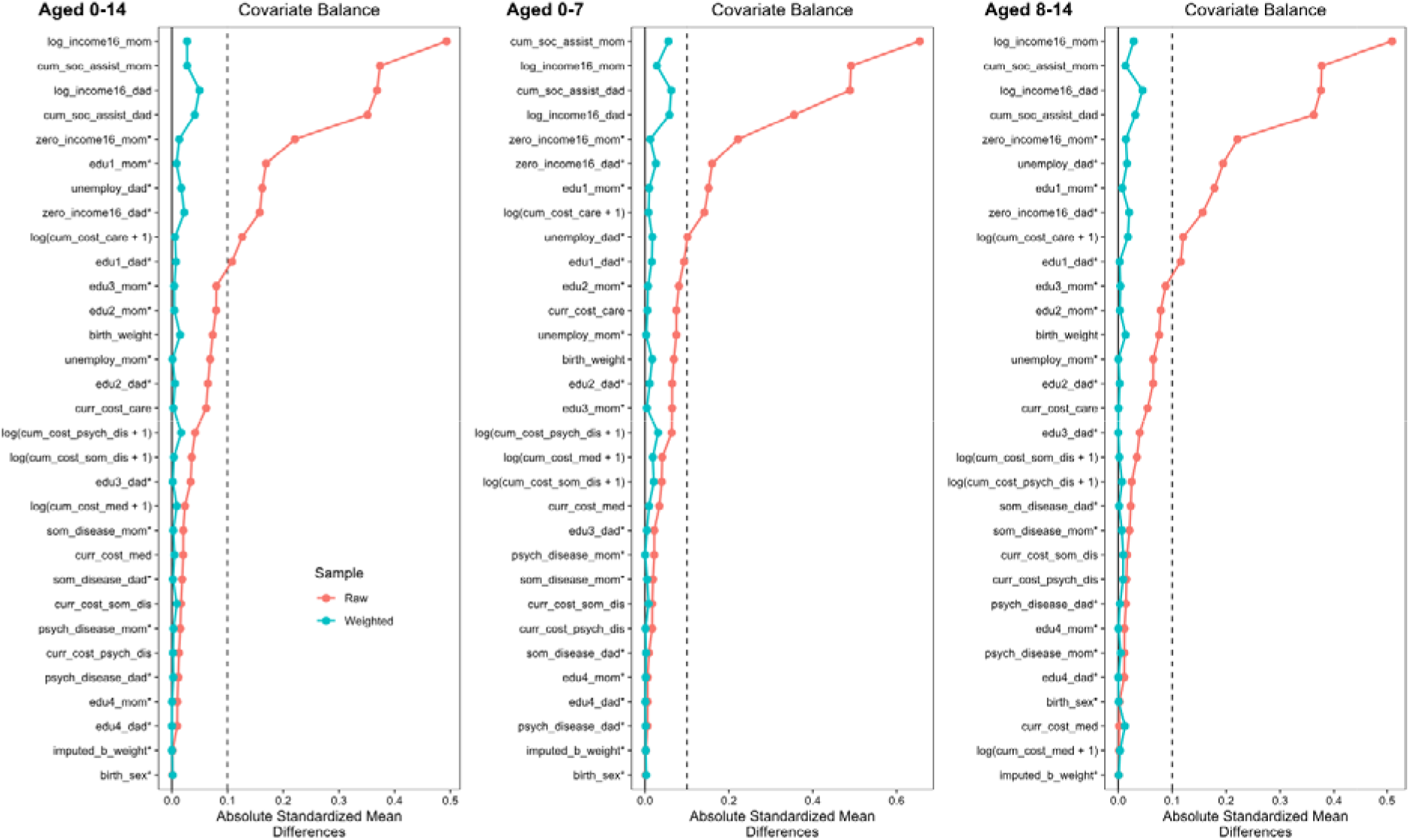
Balance characteristics of the study population in the continued social assistance entry trial. Absolute mean differences before and after weighting. Observations = 71 131. The Finnish Birth Cohort 1997.

## Ethical approval

The ethical approval for the use of this dataset has been obtained from the Ethics Committee of the Finnish Institute for Health and Welfare, THL (§572/2013).

## Data availability

The analysed data are not publicly available due to strict privacy laws. Inquiries about the access to the data can be send to Markus Keski-Säntti

